# Digoxin is Associated with Increased Mortality in Patients with Atrial Fibrillation without Concomitant Heart Failure

**DOI:** 10.1101/2020.08.07.20169888

**Authors:** Maciej Tysarowski, Rafael Nigri, Brijesh Patel, Giselle A. Suero-Abreu, Balaji Pratap, Joseph Bastawrose, Joshua Aziz, Hyoeun Kim, Eyal Herzog, Emad F. Aziz

## Abstract

**Introduction:** Atrial fibrillation (AF) is the most common arrhythmia encountered in clinical practice and is a significant risk factor for ischemic stroke and death. Digitalis has been used for more than 200 years to treat heart conditions, including AF, and its use remains controversial due to uncertain long-term morbidity and mortality.

**Methods:** We conducted a cohort study of hospitalized patients with AF assessing the effects of digoxin on longterm all-cause mortality. Patients were divided into two groups: with and without heart failure (HF). We performed multivariable Cox regression analysis to assess hazard ratios (HR) for all-cause mortality depending on digoxin treatment and used propensity score matching to adjust for differences in background characteristics between treatment groups.

**Results:** Among 2179 consecutive patients hospitalized with AF, the median age was 73 ± 14, and 52.5% of patients were male, 49% had HF, and 18.8% were discharged on digoxin. Median left ventricular ejection fraction in the whole cohort was 60 (IQR 40-65). Among patients with HF, 34.5% had preserved, 17.3% had mid-range and 48.1% had reduced left ventricular ejection fraction. The mean follow-up time was 3 ± 2.05 years. In patients without HF there was a statistically significant increased mortality in the digoxin subgroup after propensity score matching (HR = 2.23, 95% CI 1.42-3.51, p < 0.001). In contrast, in patients with HF, there was no difference in mortality between the treatment groups (p = 0.92).

**Conclusions:** Digoxin use in our study was associated with increased mortality in patients with AF and without concomitant HF.

## Introduction

Atrial fibrillation (AF) is the most common arrhythmia encountered in clinical practice, and its prevalence increases with age. It is a significant risk factor for thromboembolic stroke and affects up to 9% of the population by the age of 80 years.^1,2^ Moreover, a stroke in patients with AF is associated with higher mortality, morbidity, and longer hospital stays than those without AF.^3–5^

Two pharmacologic treatment strategies exist for AF: rate and rhythm control. The AFFIRM trial compared the overall survival benefit of treating AF with rate or rhythm control strategies. The study demonstrated no mortality difference between rhythm control with antiarrhythmics and rate control with digoxin and AV nodal blockers.^6,7^ Rate control with digoxin, beta-blockers, and calcium channel blockers is now a more utilized approach for AF treatment.

Amongst the rate control drugs, digoxin is the most controversial. ^8,9^. It is recommended for rate control in AF, by both the European Society of Cardiology and the American College of Cardiology/American Heart Association (ACC/AHA), as a second-line medication in patients with heart failure (HF) and third line in patients without HF.^10^ It has also been shown to decrease hospitalizations in patients with HF. ^11^

There is conflicting data regarding digoxin effect on all-cause mortality in patients with AF with and without concomitant HF. Some studies emphasize an association between higher all-cause mortality and digoxin use in patients with and without HF^9^, while others deny that correlation.^12^ Moreover, the follow-up in most of these studies was short, consisting of 1 year.

Our study aims to assess the effect of digoxin treatment on long term all-cause mortality in patients with AF.

## Methods

### 2.1 Study population and data acquisition

The study cohort is derived from the ACAP-RACE AF13 database that was established at St. Luke’s-Roosevelt Hospital Center in 2004 to follow all patients admitted with AF. It included all 2179 consecutive patients admitted to the institution with the diagnosis of AF or atrial flutter between September 2006 and April 2014. The comprehensive registry included all patients’ laboratory and imaging findings, and admission and discharge medications. Management of all patients was guided by utilizing the RACE pathway (R: rate control, A: anticoagulation, C: Cardioversion and E: electrophysiology) that was initially published in 2005 and updated in 2017.^14,15^ The pathway was implemented in the institution in 2005 and according to the pathway all patients were treated with either rate or rhythm control. Rhythm control with catheter ablation was offered to patients who met the inclusion criteria at that time to undergo the procedure. Mortality was confirmed by review of medical records, death certificates or the social security death index. Patients or relatives were interviewed at least twice during the follow-up period (physician-directed, scripted telephone interview). HF group was defined as having previously diagnosed HF (including HF with preserved EF), diagnosed with HF during hospitalization or having LVEF ≤35% on echocardiography during hospitalization.

### 2.2 Statistical methods

We divided all patients with AF into two groups, those with HF and those without HF and further into subgroups according to digoxin therapy (Figure 1). After verifying proportional hazard assumptions, we calculated hazard ratios (HR) for all-cause mortality in patients treated and not treated with digoxin, using Cox regression analysis. In addition, we used propensity score matching to balance the groups of patients treated, and not treated with digoxin. We used the nearest neighbor matching method with matching ratio of 1:2. Variables included in propensity score model are presented in Table 1.

**Figure 1.**
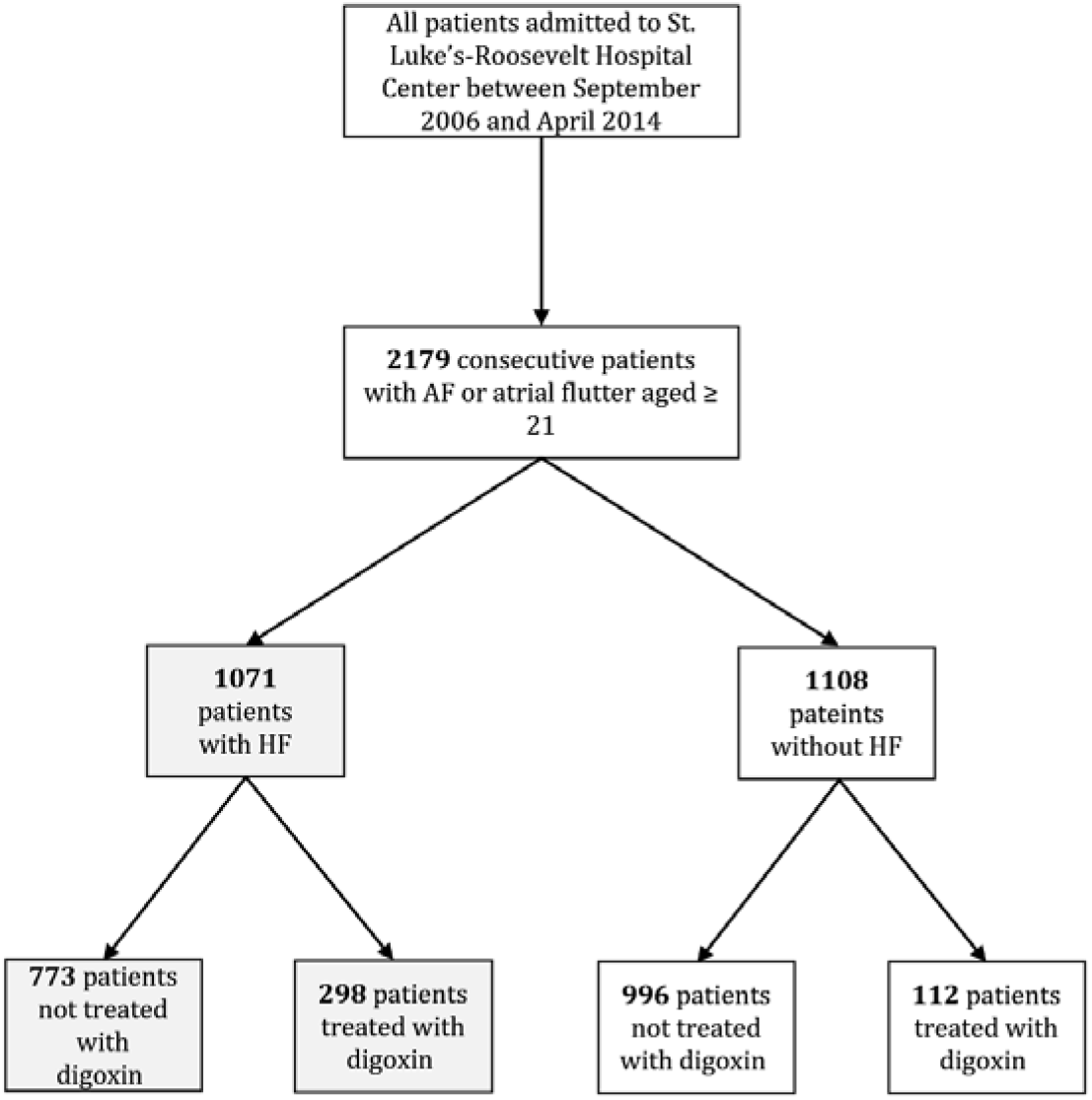
Study flow chart. Abbreviations: AF = atrial fibrillation, HF = heart failure outcome

**Table 1.**
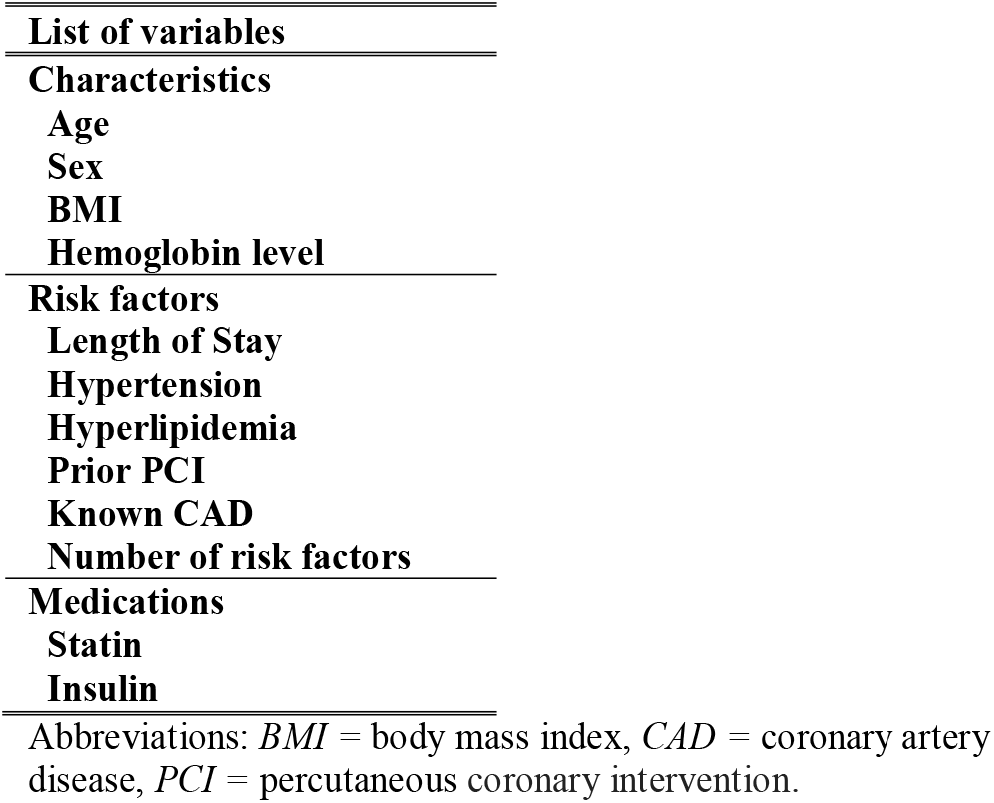
Variables included in propensity score model

Data are presented as mean ± standard deviation for normally distributed continuous variables or median (25th and 75th percentiles) for non-normal continuous variables. Shapiro-Wilk test was used to check normality of all continuous variables. Categorical variables are presented as a number (percentage). All statistical tests were 2-sided and significance was established as α= 0.05. R version 3.6.1 (R Foundation for Statistical Computing, Vienna, Austria) was used for all statistical analyses.

### 2.3 Ethics

The study met the requirements of the Declaration of Helsinki. All patients provided written follow up consent. The study protocol was accepted by Mount Sinai St. Luke’s-Mount Sinai West Hospital’s Institutional Review Board and individual consent for participation in anonymous data analysis was waived.

## Results

Unadjusted (before propensity score matching) patient characteristics are presented in Table 2. All patients had AF and/or atrial flutter. The average age was 73 ± 14, and 52.5% of patients were men. There was a high prevalence of hypertension - 77%, hyperlipidemia - 42.3%, and diabetes - 27.1%. HF was present in 49% of all patients, and 18.8% of all patients were discharged on digoxin. Median left ventricular ejection fraction in the whole cohort was 60 (IQR 40-65). Among patients with HF, 34.5% had preserved, 17.3% had mid-range and 48.1% had reduced left ventricular ejection fraction. Less than 5% of patients discharged without digoxin were taking it before the hospitalization, and for most patients discharged on digoxin, it was a new medication. The median LVEF of our study population was 60% and was significantly lower in the HF group. The mean follow-up time in our study was 3 ± 2.05 years.

Overall survival of all patients is shown in Figure 2. After adjustment (propensity score matching), there was a statistically significant increased mortality in patients treated with digoxin when compared to the non-digoxin subgroup (HR for death 1.38, 95% CI 1.09-1.76, p=0.0078). Heart failure patient survival is shown in Figure 3. After adjustment, there was no statistically significant difference in mortality between the two groups (HR for death 1.01, 95% CI 0.76-1.35, p=0.92).Survival curves for patients without HF are shown in Figure 4. According to our analysis, there was a statistically significant increase in mortality with digoxin use (HR of death 2.93, 95% CI 2.74 - 3.12). This increase in mortality was maintained after adjustment with a HR of death of 2.23 (95% CI 1.42-3.51, p<0.001) with digoxin use in this group.

**Figure 2.**
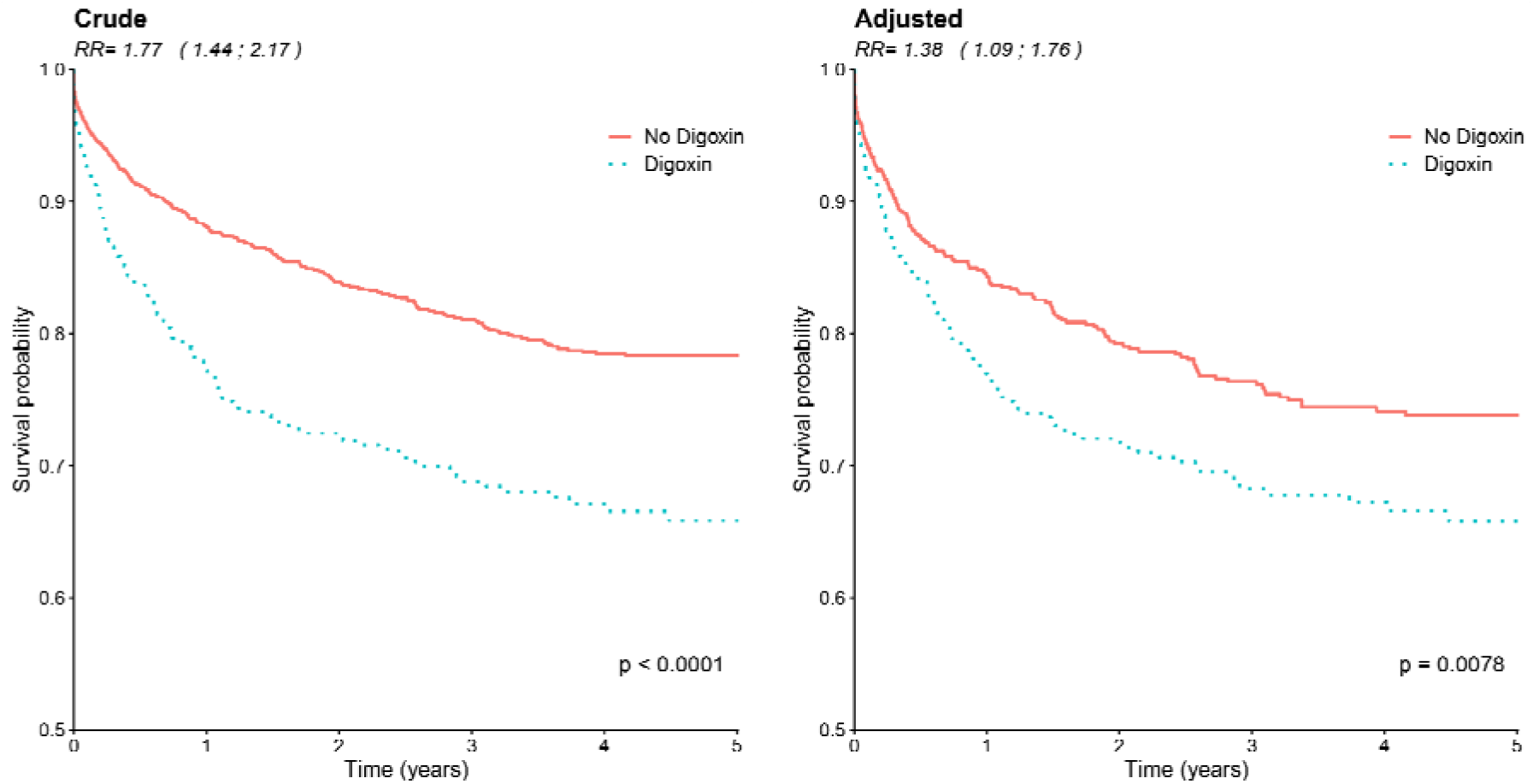
Survival probability for all patients. *Blue line* represents patients taking digoxin and *red line* those that did not use digoxin. The *left figure* shows non-adjusted survival, and the *right* shows survival after adjusting for propensity score. *RR* relative risk (95% confidence interval)

**Figure 3.**
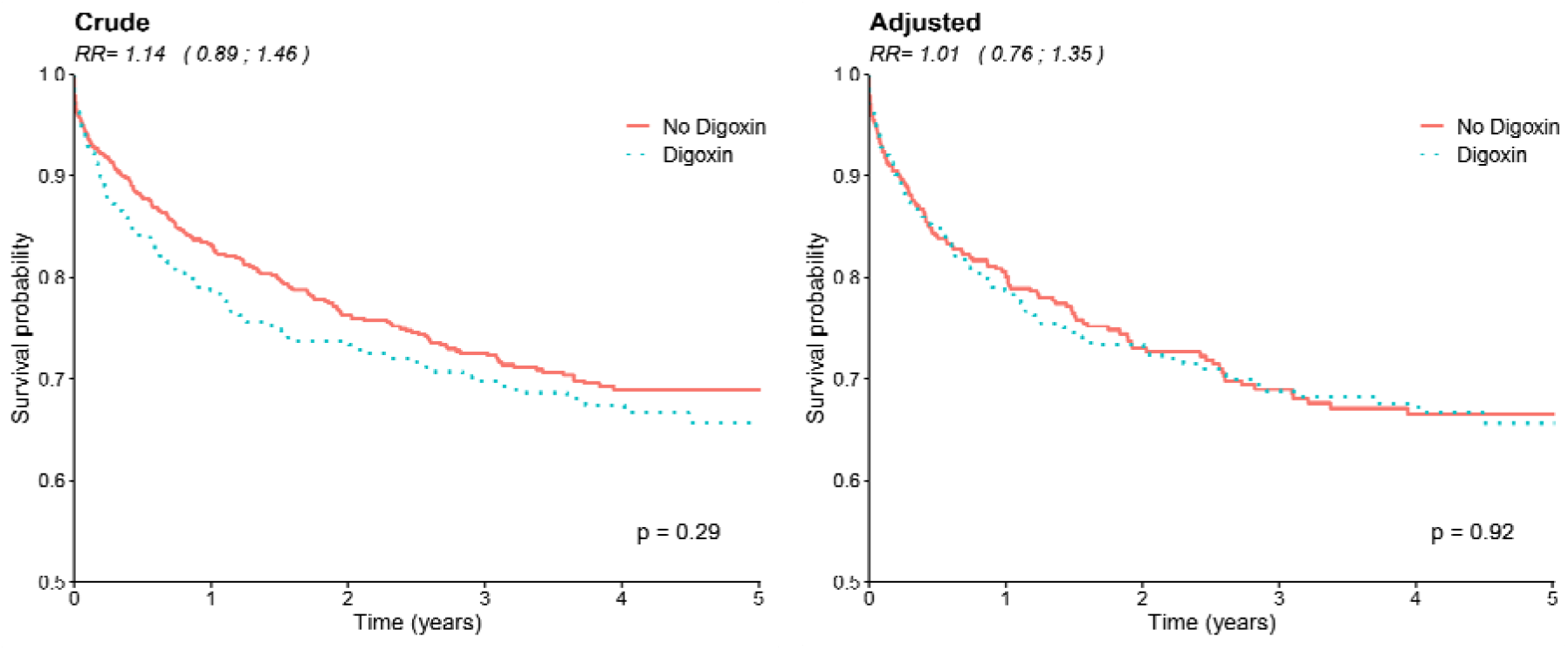
Survival probability for patients with HF. Blue line represents patients treated with digoxin and red line patients that did not use digoxin. The left figure shows non-adjusted survival, and the right shows survival adjusted for propensity score. RR relative risk (95% confidence interval)

**Figure 4.**
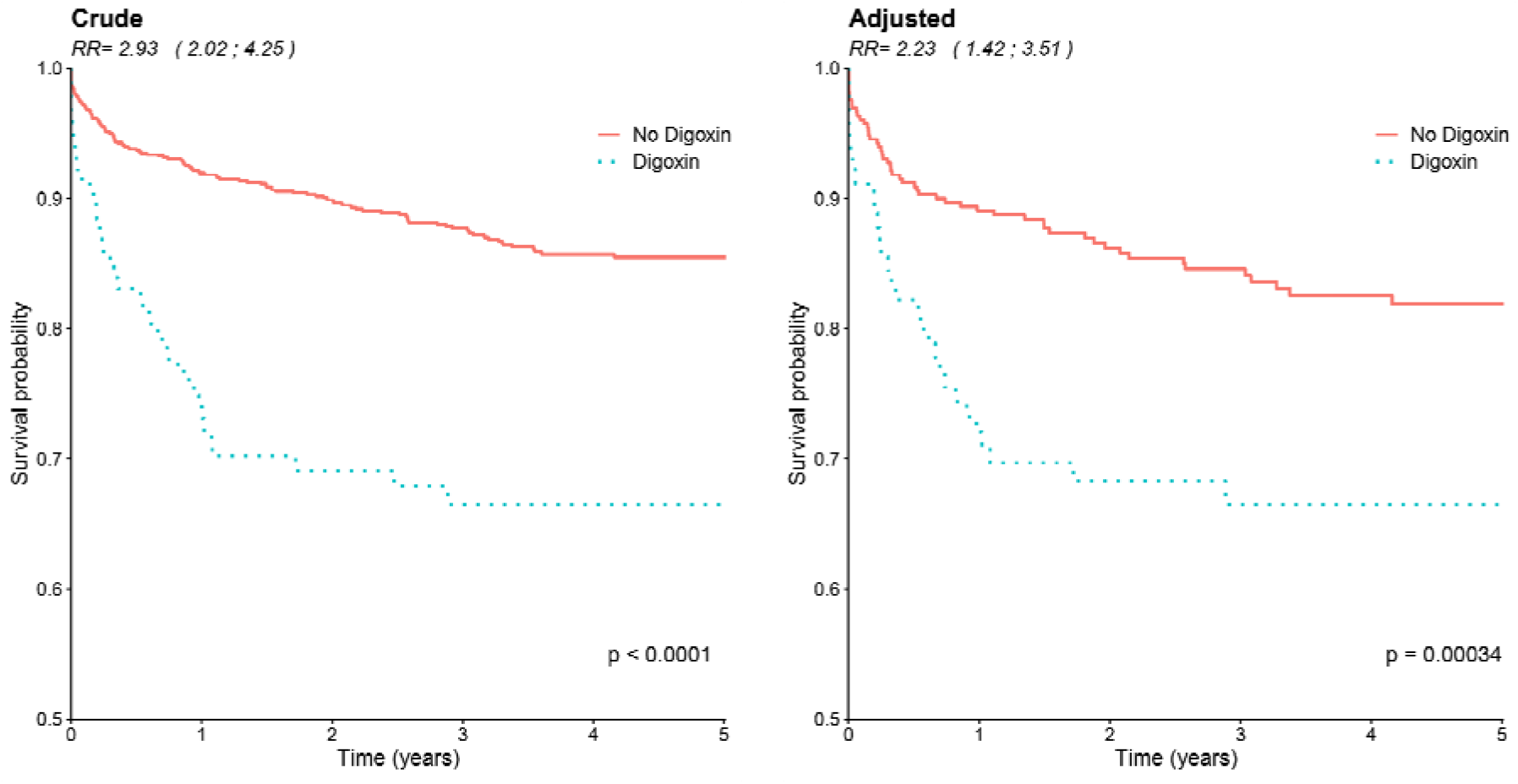
Survival probability for patients without HF. Blue line represents patients treated with digoxin and red line patients that did not use

Radiofrequency ablation rate in the whole group was 11.1% (Table 2) and was four-fold higher 14.8% in patients not treated with digoxin than the digoxin treated group 3.6%, (p=0.002). There was no statistically significant difference in RFA rates among patients with HF.

**Table 2.**
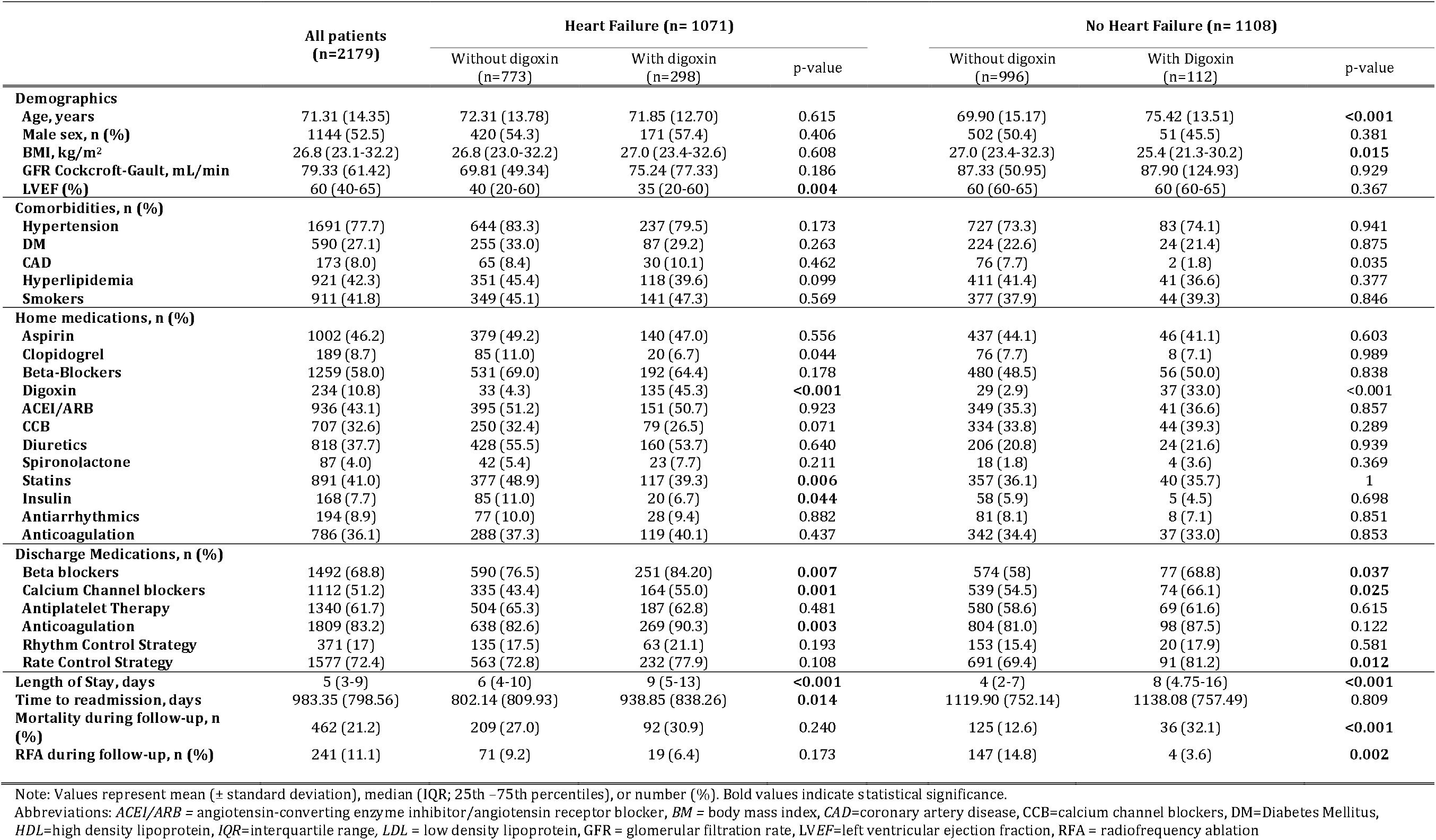
Unadjusted characteristics of study participants

|  | All patients<br>(n=2179) | Heart Failure (n= 1071) |  |  | No Heart Failure (n= 1108) |  |  |
| --- | --- | --- | --- | --- | --- | --- | --- |
|  |  | Without digoxin<br>(n=773) | With digoxin<br>(n=298) | p-value | Without digoxin<br>(n=996) | With Digoxin<br>(n=112) | p-value |
| **Demographics** |  |  |  |  |  |  |  |
| Age, years | 71.31 (14.35) | 72.31 (13.78) | 71.85 (12.70) | 0.615 | 69.90 (15.17) | 75.42 (13.51) | **<0.001** |
| Male sex, n (%) | 1144 (52.5) | 420 (54.3) | 171 (57.4) | 0.406 | 502 (50.4) | 51 (45.5) | 0.381 |
| BMI, kg/m<sup>2</sup> | 26.8 (23.1-32.2) | 26.8 (23.0-32.2) | 27.0 (23.4-32.6) | 0.608 | 27.0 (23.4-32.3) | 25.4 (21.3-30.2) | **0.015** |
| GFR Cockcroft-Gault, mL/min | 79.33 (61.42) | 69.81 (49.34) | 75.24 (77.33) | 0.186 | 87.33 (50.95) | 87.90 (124.93) | 0.929 |
| LVEF (%) | 60 (40-65) | 40 (20-60) | 35 (20-60) | **0.004** | 60 (60-65) | 60 (60-65) | 0.367 |
| **Comorbidities, n (%)** |  |  |  |  |  |  |  |
| Hypertension | 1691 (77.7) | 644 (83.3) | 237 (79.5) | 0.173 | 727 (73.3) | 83 (74.1) | 0.941 |
| DM | 590 (27.1) | 255 (33.0) | 87 (29.2) | 0.263 | 224 (22.6) | 24 (21.4) | 0.875 |
| CAD | 173 (8.0) | 65 (8.4) | 30 (10.1) | 0.462 | 76 (7.7) | 2 (1.8) | 0.035 |
| Hyperlipidemia | 921 (42.3) | 351 (45.4) | 118 (39.6) | 0.099 | 411 (41.4) | 41 (36.6) | 0.377 |
| Smokers | 911 (41.8) | 349 (45.1) | 141 (47.3) | 0.569 | 377 (37.9) | 44 (39.3) | 0.846 |
| **Home medications, n (%)** |  |  |  |  |  |  |  |
| Aspirin | 1002 (46.2) | 379 (49.2) | 140 (47.0) | 0.556 | 437 (44.1) | 46 (41.1) | 0.603 |
| Clopidogrel | 189 (8.7) | 85 (11.0) | 20 (6.7) | 0.044 | 76 (7.7) | 8 (7.1) | 0.989 |
| Beta-Blockers | 1259 (58.0) | 531 (69.0) | 192 (64.4) | 0.178 | 480 (48.5) | 56 (50.0) | 0.838 |
| Digoxin | 234 (10.8) | 33 (4.3) | 135 (45.3) | **<0.001** | 29 (2.9) | 37 (33.0) | **<0.001** |
| ACEI/ARB | 936 (43.1) | 395 (51.2) | 151 (50.7) | 0.923 | 349 (35.3) | 41 (36.6) | 0.857 |
| CCB | 707 (32.6) | 250 (32.4) | 79 (26.5) | 0.071 | 334 (33.8) | 44 (39.3) | 0.289 |
| Diuretics | 818 (37.7) | 428 (55.5) | 160 (53.7) | 0.640 | 206 (20.8) | 24 (21.6) | 0.939 |
| Spironolactone | 87 (4.0) | 42 (5.4) | 23 (7.7) | 0.211 | 18 (1.8) | 4 (3.6) | 0.369 |
| Statins | 891 (41.0) | 377 (48.9) | 117 (39.3) | **0.006** | 357 (36.1) | 40 (35.7) | 1 |
| Insulin | 168 (7.7) | 85 (11.0) | 20 (6.7) | **0.044** | 58 (5.9) | 5 (4.5) | 0.698 |
| Antiarrhythmics | 194 (8.9) | 77 (10.0) | 28 (9.4) | 0.882 | 81 (8.1) | 8 (7.1) | 0.851 |
| Anticoagulation | 786 (36.1) | 288 (37.3) | 119 (40.1) | 0.437 | 342 (34.4) | 37 (33.0) | 0.853 |
| **Discharge Medications, n (%)** |  |  |  |  |  |  |  |
| Beta blockers | 1492 (68.8) | 590 (76.5) | 251 (84.20) | **0.007** | 574 (58) | 77 (68.8) | **0.037** |
| Calcium Channel blockers | 1112 (51.2) | 335 (43.4) | 164 (55.0) | **0.001** | 539 (54.5) | 74 (66.1) | **0.025** |
| Antiplatelet Therapy | 1340 (61.7) | 504 (65.3) | 187 (62.8) | 0.481 | 580 (58.6) | 69 (61.6) | 0.615 |
| Anticoagulation | 1809 (83.2) | 638 (82.6) | 269 (90.3) | **0.003** | 804 (81.0) | 98 (87.5) | 0.122 |
| Rhythm Control Strategy | 371 (17) | 135 (17.5) | 63 (21.1) | 0.193 | 153 (15.4) | 20 (17.9) | 0.581 |
| Rate Control Strategy | 1577 (72.4) | 563 (72.8) | 232 (77.9) | 0.108 | 691 (69.4) | 91 (81.2) | **0.012** |
| Length of Stay, days | 5 (3-9) | 6 (4-10) | 9 (5-13) | **<0.001** | 4 (2-7) | 8 (4.75-16) | **<0.001** |
| Time to readmission, days | 983.35 (798.56) | 802.14 (809.93) | 938.85 (838.26) | **0.014** | 1119.90 (752.14) | 1138.08 (757.49) | 0.809 |
| Mortality during follow-up, n (%) | 462 (21.2) | 209 (27.0) | 92 (30.9) | 0.240 | 125 (12.6) | 36 (32.1) | **<0.001** |
| RFA during follow-up, n (%) | 241 (11.1) | 71 (9.2) | 19 (6.4) | 0.173 | 147 (14.8) | 4 (3.6) | **0.002** |

Note: Values represent mean ( ± standard deviation), median (IQR; 25th – 75th percentiles), or number (%). Bold values indicate statistical significance.
Abbreviations: ACEI/ARB = angiotensin-converting enzyme inhibitor/angiotensin receptor blocker, BM = body mass index, CAD=coronary artery disease, CCB=calcium channel blockers, DM=Diabetes Mellitus, HDL=high density lipoprotein, IQR=interquartile range, LDL = low density lipoprotein, GFR = glomerular filtration rate, LVEF=left ventricular ejection fraction, RFA = radiofrequency ablation

## Discussion

Our results suggest that while the use of digoxin was not associated with all-cause mortality in patients with AF and HF, it was associated with significantly increased mortality in the group of patients with AF and without HF. These results are consistent with some but not all previous studies evaluating this relationship.

### 4.1 Patients not stratified in HF and non-HF groups

In a metanalysis from 2019, Vamos et al. analyzed 37 trials evaluating the effect of digoxin on patients with AF or HF as the inclusion criterium.^16^ In the AF subgroup of 627,620 patients, which also included patients with HF, treatment with digoxin was associated with increased mortality after adjustment (HR=1.23). In comparison, our study revealed a similar adjusted HR of 1.38 for all AF patients in our cohort. The mentioned metanalysis did not stratify patients into HF and non-HF subgroups.

Two recent post hoc analyses of the AFFIRM trial have been at odds regarding the mortality associated with Digoxin.^9,17^ The propensity-adjusted analysis by Whitbeck et al. of the AFFIRM trial has found that the use of Digoxin is associated with increased mortality in AF with or without HF. These results confirmed the findings of an earlier post hoc analysis of the AFFIRM trial in which Digoxin was also associated with higher all-cause mortality. However, Gheorghiade et al. conducted a post hoc analysis of the AFFIRM trial and found no difference in mortality associated with Digoxin use in AF after propensity matching. In the latter analysis in order to eliminate intention bias digoxin was not considered as a time-dependent treatment variable and 1352 patients were excluded. These excluded patients had higher unadjusted mortality than those included in the analysis. While the findings were consistent with the AFFIRM trial results, the exclusion of a large cohort of patients with a high mortality limits the external validity of the study.

### 4.2 Patients without HF

A 2015 metanalysis of 12 studies that used digoxin primarily for rate control of AF concluded that digoxin is associated with increased all-cause mortality in patients without HF with HR of 1.38 (95% CI, 1.12-1.71).^18^ ATRIA-CVRN is a cohort study consisting of over twenty-three thousand patients with newly diagnosed AF with no history of HF. It demonstrated a significant increase in mortality with digoxin (HR 1.71; 95% CI, 1.52-1.71).^19^ An analysis using the RIKS-HIA database with over 4000 patients discharged from the Coronary Unit in Sweden compared outcomes of patients with AF, HF, or both. Digoxin was demonstrated again to have an increased 1-year overall mortality in patients without HF (HR=1.42; 95% CI, 1.17-1.53).^20^ These three studies had similar results, but with a lower hazard ratio when compared to our analysis (HR=2.23). One of the main differences between the RIKS-HIA analysis and our study is the fact that our study included all patients admitted to the hospital, not only those admitted in the coronary care unit (CCU).

### 4.3 Patients with HF

In the mentioned metanalysis by Chamaria et al., researchers concluded that in patients with AF and HF, digoxin was not associated with increased mortality (HR 1.08, 95% CI 0.99–1.18).^18^ In another subgroup of patients from the RIKS-HIA database that comprised of patients with both AF and CHF, there was no statistically significant difference in mortality between patients who were discharged with or without Digoxin (RR 1.00, 95% CI, 0.94-1.06).^20^ Analysis of our cohort concurs with these two reports, no statistical difference in mortality was found between the patients with HF treated with or without digoxin. In patients with HF in addition to AF, digoxin could also be of benefit due to its neurohormonal action and effect in decreasing chronotropy while increasing inotropy.^21^

In patients with AF and HF the Serum Drug Concentration (SDC) has directly affected the efficacy and associated mortality of the drug.^22,23^ The DIG trial is the only randomized clinical trial that assessed the mortality and morbidity of Digoxin in patients with HF. The study revealed no difference in mortality and a decrease in hospital admission with digoxin use.11 Our results are in agreement with these findings; we found no difference in mortality but a statistically longer time to readmission in the digoxin subgroup. Other studies, including a post hoc analysis of the DIG trial, demonstrated increased mortality with a SDC is greater than 1 ng/ml and a trend towards decreased mortality when the SDC is between 0.5 and 0.9 ng/ml.^22,23^ Although this appears to be a discrepancy between the DIG trial and the post hoc analysis, DIG trial reported that the average SDC at one month in the Digoxin arm was less than 0.9 ng/ml. A SDC of less than 1 ng/ml has also been shown to have beneficial hemodynamic, neurohormonal, and clinical effects.^24,25^ These findings are also corroborated by a post hoc analysis of the ARISTOTLE trial. A Digoxin level equal or above 1.2 ng/ml was independently associated with an increased risk of death and sudden death in patients with AF with or without HF.^26^ Recent analysis of the AF-CHF trial contradicts the conclusion that digoxin is not associated with increased mortality in patients with HF and AF. The analysis revealed a high risk of death among all patients, but had a statistically significant higher all cause deaths, cardiac deaths and arrhythmic deaths in the Digoxin subgroup. These findings could have been influenced by the Digoxin serum level which was not measured in the study. ^27^

### 4.4 Rates of radiofrequency ablation

In the non-HF group, the rate of radiofrequency ablation was more than four times higher in the group discharged without digoxin, a statistically significant difference. This may suggest that digoxin could potentially provide substantial AF symptom improvement secondary to better rate control and decreasing the need for invasive procedures. However, during the study duration the results of the CASTLE-AF was not yet published and there might have been procrastination in offering ablation to patients with severe heart failure.^28^

#### Study limitations and strengths

We conducted an observational and single-center study. Nonrandom assignment to digoxin treatment in study participants is reflected in differences between groups in Table 2. There are reports suggesting that prescription bias in observational digoxin studies far exceeds the true effect of the treatment, and thus, even after adjustment, they fail to capture the real consequences of the medication.29 Since digoxin would have been added as 3rd line agent in patients without HF, it can be assumed that rate control was difficult and these patients would have had higher mortality regardless of digoxin use in these patients. Our registry contained SDC levels only for a limited number of patients, mostly obtained for suspected digoxin toxicity, so we did not include them in our analysis to avoid selection bias. We also had no data on the further specification of HF into HF with preserved ejection fraction, HF with moderately reduced and reduced ejection fraction.

Our study included a relatively large population of 2197 consecutive diverse inner-city hospital patients. It had a follow-up period of mean of 3 years, longer than in most previous digoxin studies. We evaluated all-cause mortality, being one of the most clinically meaningful outcomes. We used propensity score matching to account for the digoxin prescription bias, and the differences in outcomes persisted, although with less effect size.

#### Conclusions and clinical indications

Digoxin use was associated with an increased all cause-mortality in patients with AF and without concomitant HF. No difference in mortality was established in patients with AF with HF. These results suggest the need for the careful use of digoxin, despite being recommended in both European and ACC guidelines as a third-line agent of rate control in AF in patients without HF.

There is still controversy about whether digoxin increases mortality in patients with AF, especially in the subgroup without HF. This needs to be studied further with large randomized clinical trials.

#### CRediT authorship contribution statement

**Maciej Tysarowski:** conceptualization, methodology, investigation, validation, data curation, writing - original draft, review & editing, visualization, project administration, formal analysis, software, resources. **Rafael Nigri:** conceptualization, methodology, investigation, validation, data curation, writing - original draft, review & editing, visualization, project administration, formal analysis, software, resources. **Brijesh Patel**: investigation, resources, data curation, writing - review & editing. **Giselle A. Suero-Abreu**: investigation, resources, data curation, writing - review & editing. **Balaji Pratap**: investigation, resources, data curation, conceptualization, methodology. **Joseph Bastawrose**: investigation, resources, data curation,, methodology. **Joshua Aziz**: data curation, methodology. **Hyoeun Kim**: investigation, resources, data curation, conceptualization, methodology. **Eyal Herzog**: investigation, resources, data curation, conceptualization, methodology. **Emad F. Aziz**: conceptualization, methodology, investigation, validation, resources, data curation, writing - original draft, review & editing, visualization, project administration.

## Data Availability

not available

